# E-cigarette market share by nicotine claims

**DOI:** 10.1101/2024.08.14.24311761

**Authors:** Shaoying Ma, Marielle C. Brinkman, Micah Berman, Theodore Wagener, Ce Shang

**Author notes:** **Corresponding author:** Shaoying Ma, Ph.D., The Ohio State University Center for Tobacco Research, 3650 Olentangy River Road Suite 110, Columbus OH 43214, USA.

## Abstract

**Background:** Nicotine forms (salt vs. freebase) and isomers (synthetic vs. tobacco-derived) are key characteristics of e-cigarettes that manufacturers manipulate, and “tobacco-free” claims may have served to attract new consumers and increase their intention to use.

**Method:** This study presents a snapshot of nicotine marketing claims for e-cigarettes using Nielsen ScanTrack data from brick-and-mortar stores. Market share was calculated as the ratio of unit sales of each nicotine claim category to the total unit sales of e-cigarettes during the four weeks ending 1-20-2024.

**Results:** We summarized the market share for the following six nicotine form/isomer category: 1) nicotine (77%), 2) nicotine salt (10%), 3) synthetic nicotine (2%), 4) zero tobacco or tobacco-free (2%), 5) zero nicotine or nicotine-free (0.03%), and 6) no claim or CBD/hemp/cannabis (9%).

**Conclusion:** The market share of products that explicitly carried nicotine salt claims (10%) or synthetic nicotine or tobacco-free claims (2% each) was notable. This study informs regulatory authorities on the recent trend of nicotine claims marketed by the e-cigarette industry, which may be contributing to the use of these products or addiction to nicotine among young people and non-users.

---

Nicotine dimensions including forms (salt vs. freebase) and isomer (synthetic vs. tobacco-derived) are key characteristics of e-cigarettes that manufacturers manipulate to influence product addictiveness.^1–6^ As e-cigarettes have become the most popularly used tobacco product by young people, “tobacco-free” claims about synthetic nicotine products marketed by the industry may have served to attract new consumers, increase their intention to use, and reduce their harm perception about e-cigarettes.^7^ “Tobacco-free” could possibly be seen as an unauthorized modified risk tobacco product (MRTP) claim since it decreases people’s perceived relative harm of e-cigarettes to other tobacco products.^8^ Alternatively, nicotine claims may also serve to encourage adult smokers to switch to e-cigarettes, likely a less harmful tobacco product.

Leading brands of e-cigarettes such as JUUL and Puff Bar use high concentrations of nicotine salts, a form of nicotine that is less harsh on the throat.^9^ Initiating nicotine use with high concentrations of palatable nicotine may lead to a greater likelihood of long-term use of nicotine and tobacco products.^5,10^ Furthermore, synthetic nicotine is now an e-liquid ingredient that is just as economically feasible as tobacco-derived nicotine, and is often marketed with the “tobacco-free” claim, wording that may reduce the perceived harm or addictiveness of the products among consumers.^11,12^ Synthetic nicotine can specify something about the nicotine isomer that is used in the product. There are two primary nicotine isomers: (R)-nicotine, and (S)-nicotine. Tobacco-derived nicotine contains > 99% S-nicotine that has been extracted from tobacco plants, whereas synthetic nicotine is prepared from precursor chemicals and could be either > 99% S-nicotine or a racemic mixture of S- and R-nicotine isomers.^2^ Racemic nicotine mixtures may have a different level of addiction liability compared to those containing > 99% S-nicotine.^11^

Because manufacturers’ nicotine claims may affect consumer use, it is crucial to monitor the retail sales of e-cigarette products with different claims of nicotine dimensions. This study presents a 4-week snapshot of the primary nicotine marketing claims for e-cigarette products based on Nielsen ScanTrack data, which provides information about retail sales in brick-and-mortar stores, i.e., pharmacies, convenience stores, and gas stations (monitoring period ending 1-20-2024). **Figure 1** shows a summary of the market share and number of products by the following six nicotine-related claim categories: nicotine, nicotine salt, synthetic nicotine, zero tobacco (i.e., tobacco-free), and zero nicotine (i.e., nicotine-free), and other (i.e., no nicotine-related claims, or CBD/hemp/cannabis). Market share was calculated as the ratio of unit sales of each category to the total unit sales of e-cigarettes during the four-weeks period. The results are presented as the following: 1) nicotine (1,228 unique products, 77% market share), 2) nicotine salt (959 unique products, 10% market share), 3) synthetic nicotine (108 unique products, 2%), 4) zero tobacco (96 unique products, 2%), 5) zero nicotine (12 unique products, 0.03%), and 6) other (686 unique products, 9%).

**Figure 1.**
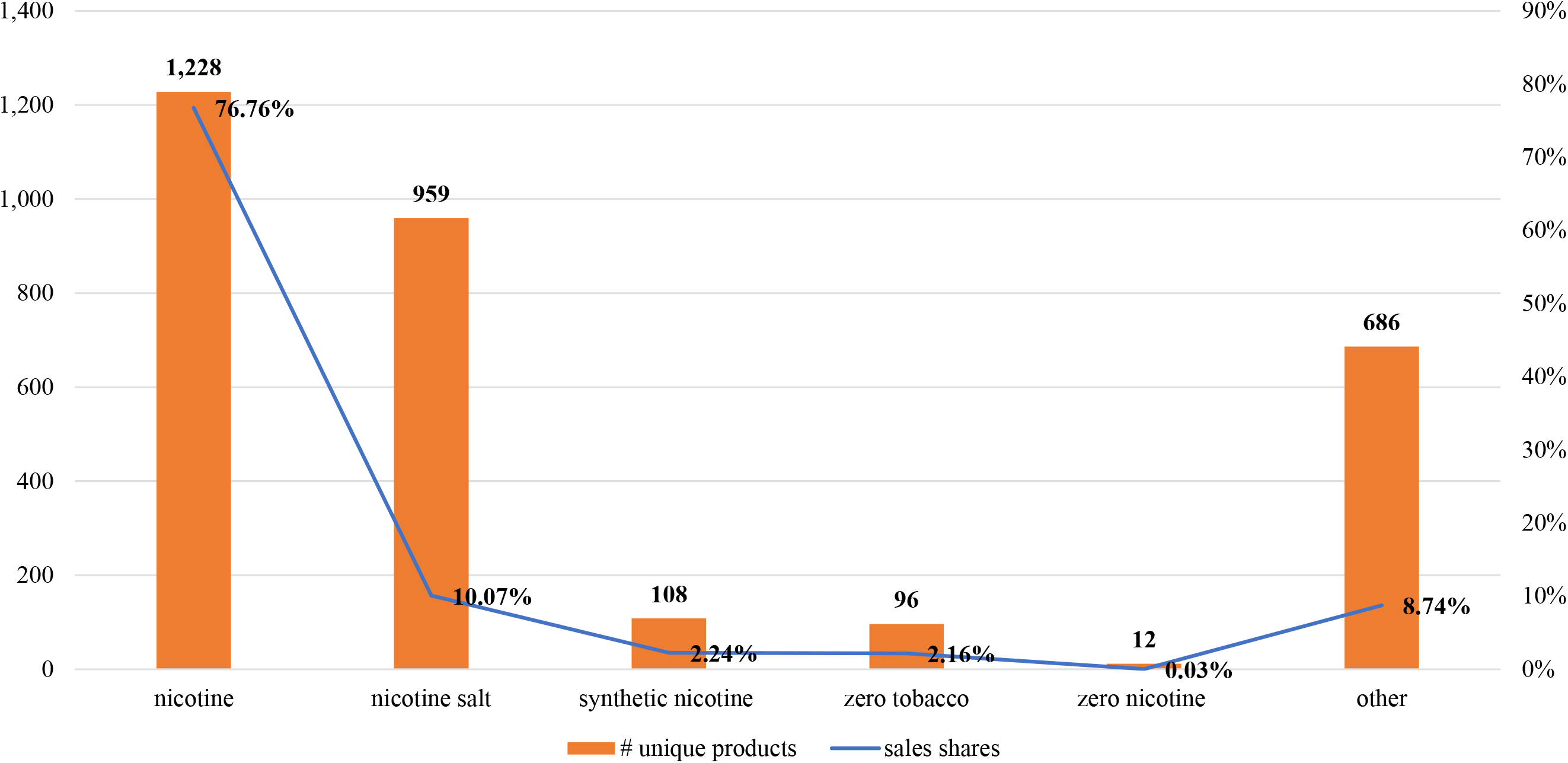
E-cigarette market shares by nicotine claims, 4 weeks ending 1/20/2024. Note: “Other” is none (638, 8.70%) or CBD/hemp/cannabis (48, 0.04%) combined

These findings show that while the vast majority of e-cigarette products marketed themselves as containing nicotine (also disclosed on their required warning labels), the market share of products that explicitly carried nicotine salt claims was substantial (10%) and the market share of products making synthetic nicotine or tobacco free claims was also notable (2% each). Thus, nicotine salt, synthetic tobacco, and tobacco free claims may be contributing to the use of these products. Future research should investigate how nicotine-related claims may intersect with product characteristics to influence e-cigarette harm perceptions and use intentions of different consumers.^13-15^

## Data Availability

The data that support the findings of this study are available from Nielsen. Restrictions apply to the availability of these data, which were used under license for this study.

## Acknowledgement

The authors’ own analyses and calculations based in part on data reported by Nielsen through its contract service for the SMOKELESS TOBACCO and ALTERNATIVE VAPOR category for the 4-week period ending 2024, for the total US market and FSMG+ Convenience channel (Copyright © 2024, Nielsen Consumer LLC). The conclusions drawn from the Nielsen data are those of the authors and do not reflect the views of Nielsen. Nielsen is not responsible for and had no role in was not involved in analyzing and preparing the results reported herein.

